# Integrating Prior Qualitative Coronary Artery Calcium Assessment into an Established Chest Pain Algorithm: Effect on Risk Stratification Outcomes

**DOI:** 10.64898/2025.12.02.25341520

**Authors:** Aradhya Abrol, Smith Frimpong, Rifla Hassan, Dima Khalil, Samuel Schwartzwald, Martin E. Matsumura

## Abstract

**Background:** Chest pain (CP) patients with moderate elevations of high sensitivity cardiac troponins benefit from additional risk stratification strategies to improve Emergency Department decision-making. In the present study we evaluated the impact of adding the results of opportunistic detection of coronary calcium (ODCAC) from prior routine chest computed tomography scans (CT) to both high sensitivity troponin T (hsTnT) accelerated diagnostic protocol and determination of the HEART score (HS).

**Methods:** Retrospective study of 1,525 consecutive patients evaluated for CP with moderate peak elevations of hsTnT (12-51 ng/L). Thirty-day major adverse cardiovascular events (MACE) were assessed based on peak hsTnT, HS, and ODCAC on chest CT performed within 5 years of CP presentation. Firth’s penalized logistic regression was employed to assess the combined predictive power of these variables.

**Results:** The median age of the cohort was 69, 59% were male, and 81% had ODCAC on review of chest CT. Patients with MACE had higher peak hsTnT, median HS, and higher proportion of ODCAC+ versus those without MACE. Absence of ODCAC demonstrated a negative predictive value of 99.1% for excluding MACE. A logistic regression model including peak hsTnT, HS and ODCAC was successfully fitted (model p=0.002) and revealed that peak hsTnT was the strongest predictor of MACE (OR 1.05 per unit increase, 95% CI 1.02-1.09, p=0.005). A HEART score >3 had borderline-significant association (OR 14.71, 95% CI 0.88-246.19, p=0.061). In this comprehensive model the direct statistical effect of ODCAC was attenuated (p=0.111).

**Conclusion:** The presence of ODCAC was a predictor of and its absence yielded a high negative predictive value for MACE. However, ODCAC did not add incremental risk stratification to a risk algorithm including hsTnT and HS. The lack of additive prognostic value of ODCAC in this particular cohort may be related to the advanced age and resultant high background rate of CAC.

## Introduction

Chest pain (CP) remains one of the most common presenting complaints in emergency departments (ED) worldwide, accounting for over 8 million visits annually in the United States alone (1). The challenge of accurately identifying patients at risk for major adverse cardiac events (MACE) while avoiding unnecessary hospitalizations continues to be a cornerstone of emergency cardiovascular care. High-sensitivity cardiac troponin assays have revolutionized the approach to acute coronary syndrome diagnosis, allowing in some instances for more efficient risk stratification of CP (2,3). However, a significant population of patients presents with moderately elevated high-sensitivity troponin levels that fall approximately between the 99th percentile upper reference limit and what is traditionally considered a “positive” result. These patients represent a diagnostic challenge, as they have an intermediate risk for adverse outcomes that requires careful risk stratification to guide appropriate disposition and treatment decisions (4,5). The HEART score (HS) has emerged as a validated clinical decision tool that combines clinical history, electrocardiographic findings, age, risk factors, and troponin levels to stratify patients into low, intermediate, and high-risk categories (6,7). Recently, its role in augmenting decision-making based on high sensitivity troponins has been demonstrated (8).

Coronary artery calcium (CAC) scoring has been established as a powerful predictor of cardiovascular events in asymptomatic populations and has been incorporated into primary prevention guidelines (9,10). The presence of coronary calcium reflects the burden of coronary atherosclerosis and has been shown to provide incremental prognostic information beyond traditional risk factors. While quantitative CAC scoring using dedicated cardiac CT protocols is the gold standard, qualitative (i.e., present/absent) assessment of coronary calcium, in particular CAC obtained from “opportunistic detection of CAC” (ODCAC) on prior routine chest CT imaging may provide similar prognostic information with the advantage of utilizing already available imaging data (11-13). Many patients presenting to the ED with chest pain have previously undergone chest CT imaging for various indications prior to the CP presentation. The ability to leverage this existing imaging data for risk stratification could provide valuable additional information without requiring additional ED evaluation time and need for on-site interpretation of both CAC and incidental CT findings, radiation exposure, or healthcare costs. Prior studies have demonstrated that ODCAC assessment on non-gated chest CT can reliably identify the presence of coronary atherosclerosis, though its incremental value in ED risk stratification algorithms and guidance remains unclear (14,15).

The objective of this study was to evaluate whether the addition of ODCAC assessment from prior routine chest CT imaging provides incremental risk stratification value beyond high sensitivity troponin T (hsTnT) levels and HS in patients presenting to the ED with CP and moderately elevated troponin levels.

## Methods

The data that support the findings of this study are available from the corresponding author upon reasonable request and approval of Geisinger Health System.

This retrospective cohort study was conducted in the 5 EDs of Geisinger Health System, a tertiary-care academic center located in northeastern and central Pennsylvania. The study was approved by the institutional review board with a waiver of informed consent due to its retrospective design. The study cohort consisted of consecutive patients who presented to the ED with a primary diagnosis of CP from May 2022 to August 2023 and had application of the CP accelerated diagnostic protocol with moderate elevations in hsTnT levels (defined as a peak measure 12–51 ng/L)(2). From this initial cohort, patient electronic records were reviewed for ODCAC data from prior chest CTs within 5 years of the index ED visit and index ED HS data.

Inclusion criteria were 1) age ≥18 years, 2) primary ICD-10 diagnosis of CP, 3) application of the CP accelerated diagnostic protocol with peak hsTnT level between 12-51 ng/L during the index visit, and 4) availability of at least one chest CT scan performed within 5 years prior to the index emergency department visit, and 5) discharge from the ED with 30-day follow up data available in the electronic medical record (EMR). The primary outcome were major adverse cardiovascular events (MACE) within 30 days of ED discharge, defined as all-cause mortality and recurrent myocardial ischemic events.

Electronic medical records were systematically reviewed to collect demographic data, clinical characteristics, laboratory values, electrocardiographic findings, and imaging results. HS components were calculated retrospectively using standardized definitions as previously described (6). The HEART score incorporates five elements: History (slightly suspicious = 0 points, moderately suspicious = 1 point, highly suspicious = 2 points), ECG (normal = 0, non-specific repolarization disturbance = 1, significant ST deviation = 2), Age (<45 years = 0, 45-65 years = 1, >65 years = 2), Risk factors (no risk factors = 0, 1-2 risk factors = 1, ≥3 risk factors or history of atherosclerotic disease = 2), and Troponin (≤normal limit = 0, 1-3x normal limit = 1, >3x normal limit = 2). The HS calculation utilizing a previously described EMR-embedded best-practice alert and the hsTnT accelerated diagnostic protocol applied to the included patients have been previously described (8).

Non-gated chest CT imaging obtained for various clinical indications within the 5-year period preceding the index chest pain visit were independently reviewed by members of the study team who were trained reviewers and were blinded to clinical outcomes. The presence of ODCAC was assessed through visual inspection of epicardial coronary arteries (left main, left anterior descending, left circumflex, and right coronary artery) and was recorded using a binary classification system (i.e., ODCAC present/absent in any of the coronary arteries).

Comparisons of numerical and categorical data were made using student’s T test and chi square analysis, respectively. A multivariate logistic regression model was constructed using significant variables on univariate evaluation. Due to challenges encountered in applying standard logistic regression models to these data—including high correlation between variables (ODCAC and dichotomized HEART score) and near-perfect prediction scenarios (i.e., variables strongly predictive of outcomes)—a penalized logistic regression method, Firth’s penalized logistic regression, was utilized. Firth’s logistic regression addresses rare event occurrences, perfect prediction issues, and collinearity between strong predictors by applying a bias-reduction penalty, thereby providing more robust estimates. Logistic regression data were reported as odd ratios and 95% confidence intervals.

Statistical analyses were performed using Stata 19 (StataCorp LLC, College Station TX) and Sigmastat (Grafiti LLC, Palo Alto CA). For all comparisons, p<0.05 was considered significant.

## Results

A total of 1525 patients were included in the assessment. Routine chest CT for assessment of ODCAC was available in 1220/1525 (80.0%), with 988 (81.0%) noted to have ODCAC. A sub-cohort (n=627) had a complete dataset of hsTnT, HS, and ODCAC. Evaluation steps and patient numbers are summarized in the **Figure**.

**Figure:**
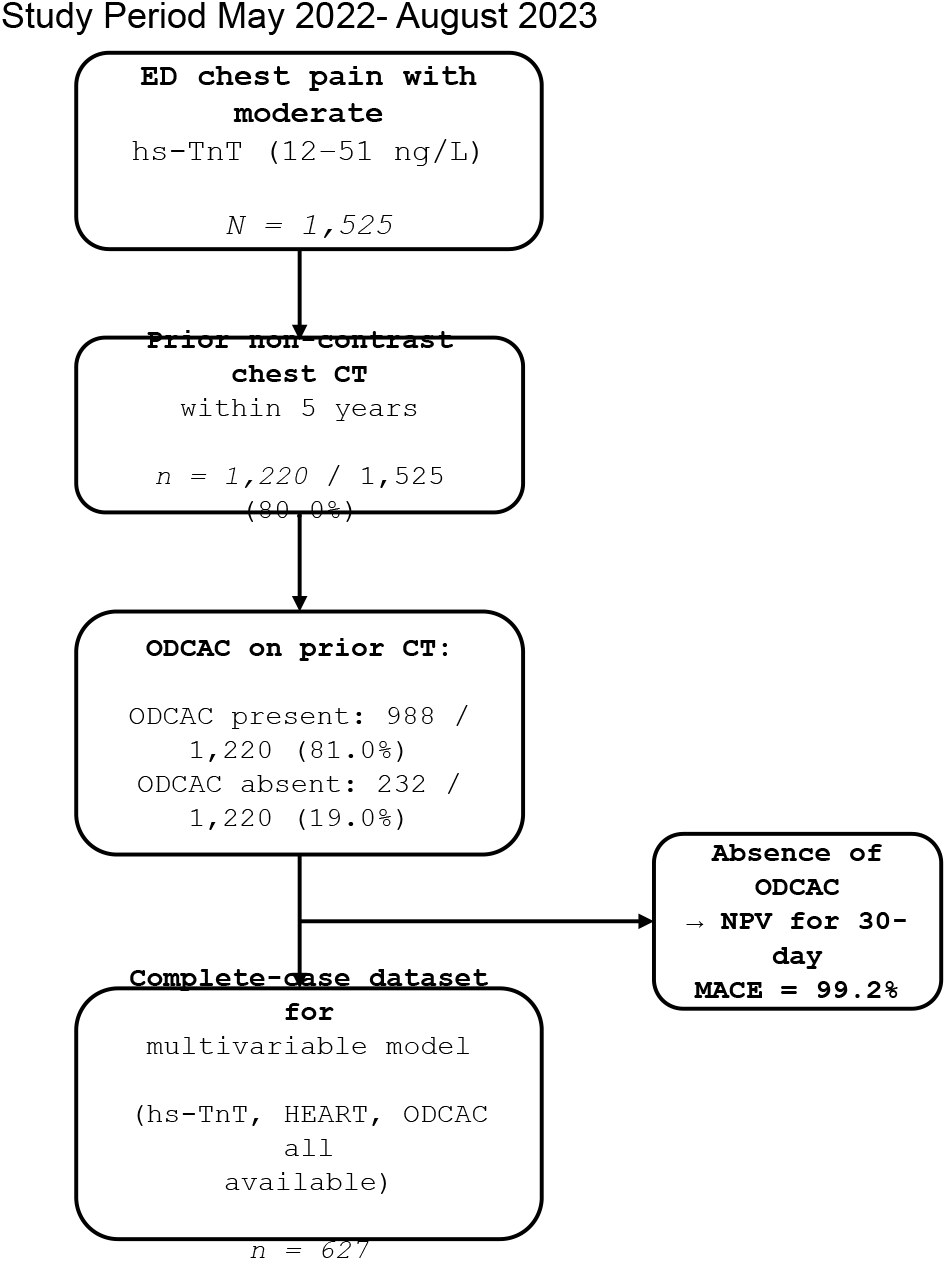
Study evaluation steps and resultant patient numbers.

Of the total cohort, 64/1525 (4.2%) suffered MACE by 30-days post-discharge. Demographics and risk stratification characteristics by MACE +/- are summarized in **Table 1**. With regard to risk stratification results, MACE(+) patients were significantly more likely to have a history of CAD vs. MACE(-) patients. MACE(+) patients had significantly higher median peak hsTnT (23ng/L vs 18ng/L, p=0.001) and median HS (5 vs. 4, p<0.001) compared to patients without 30-day MACE. Patients who experienced 30-day MACE had a higher proportion of ODCAC (96.1% vs. 80.3%, p=0.003) versus those without MACE.

**Table 1:** Patient characteristics, total and comparing MACE+/−.

|  | All patients (n-1525) | MACE+ (n=64) | MACE- (n=1461) | P (MACE+ vs MACE-) |
| --- | --- | --- | --- | --- |
| Age, median (25-75%tile) | 69 (59-79) | 68 (60-76) | 69 (59-79) | 0.97 |
| Sex, M (%) | 898 (58.9) | 43 (67.2) | 855 (58.5) | 0.21 |
| Diabetes (%) | 277 (18.2) | 7/(10.9) | 250 (17.1) | 0.26 |
| Hypertension (%) | 414 (27.1) | 19 (29.7) | 395 (27.0) | 0.74 |
| Hyperlipidemia (%) | 307 (20.1) | 13 (20.3) | 294 (20.1) | 0.90 |
| Clinical diagnosis of CAD (%) | 715 (46.9) | 46 (71.9) | 669 (45.8) | <0.001 |
| Aspirin treatment (%) | 443 (29.0) | 25 (39.1) | 418 (28.6) | 0.10 |
| Statin treatment (%) | 284 (18.6) | 16 (25.0) | 268 (18.3) | 0.24 |
| Peak hsTnT, median, ng/L (25-75%tile) | 18 (14-25) | 23 (16-31) | 18 (14-35) | 0.001 |
| Heart score, median (25-95%tile) | 4 (3-5) | 5 (4-6) | 4 (3-5) | <0.001 |
| ODCAC+ (%) | 988/1220 (80.9) | 49/51 (96.1) | 939/1169 (80.3) | 0.003 |

CAD, coronary artery disease; hsTnT, high sensitivity troponin T; ng/L, nanograms per liter; ODCAC, opportunistic detection of coronary calcium

HS and ODCAC were assessed as univariate predictors of 30-day MACE (**Table 2**). Both dichotomized HS expressed as ≤3 vs. >3 (OR=9.273, 95% CI 2.213-38.864, p=0.001) and ODCAC present vs. absent (OR=6.007, 95% CI 1.450, 24.884, p=0.007) were associated with 30-day MACE. The absence of ODCAC demonstrated a strong negative predictive value (NPV) of 99.2% and weak positive predictive value (PPV) of 5.0% for 30-day MACE.

**Table 2:** Univariate analysis of ODCAC and Heart Score as predictor of 30d MACE.

|  | Odds ratio | 95% Confidence interval | p |
| --- | --- | --- | --- |
| ODCAC +/- | 6.007 | 1.450, 24.884 | 0.007 |
| Heart Score >3/<u>≤</u>3 | 9.273 | 2.213, 38.864 | 0.001 |
| ODCAC, opportunistic detection of coronary calcium |  |  |  |

On patients with complete data set including hsTnT, CT, and HS we performed a Firth’s penalized logistic regression, assessing the combined predictive value of peak hsTnT (continuous variable), dichotomized HS (≤3 versus >3), and ODCAC presence/absence (**Table 3**). In this model peak hsTnT was the most robust predictor (OR 1.05 per unit, 95% CI: 1.02–1.09; p = 0.005). The association of ODCAC was attenuated (p = 0.111) in the full model, and the HS >3 showed a borderline association (OR = 14.71, 95% CI: 0.88–246.19; p = 0.061).

**Table 3:** Firth’s logistic regression analysis.

|  | Odds ratio | 95% Confidence interval | p |
| --- | --- | --- | --- |
| ODCAC +/- | 9.943 | 0.591,167.270 | 0.111 |
| Heart Score | 14.712 | 0.879, 246.189 | 0.061 |
| Peak hsTnT | 1.052 | 1.016, 1.090 | 0.005 |

hsTnT, high sensitivity troponin T; ODCAC, opportunistic detection of coronary calcium

## Discussion

Defining appropriate CP patients for ED discharge remains a challenge. In addition to biomarkers and bioclinical risk scores, the addition of imaging tests to augment risk stratification of CP is an attractive strategy with growing supportive evidence (3,16). However, imaging adds cost and adds time to the ED length of stay for CP assessments. The prospect of using existent imaging data such as ODCAC on prior unenhanced chest CT scans could potentially add enhanced risk stratification without additional time or cost burden (17). Prior studies have validated the identification of CAC on routine ungated chest CT vs. formal gated CAC scoring (15,18). The present study evaluated the value of adding ODCAC to a contemporary risk stratification strategy of a hsTnT accelerated diagnostic protocol and application of the HEART score. This study demonstrated that while ODCAC assessment on prior chest CT imaging was a predictor of 30-day major adverse cardiac events in chest pain patients with moderately elevated troponin levels, it did not provide significant incremental risk stratification value when added to a comprehensive model including hsTnT and HS. These findings have several important clinical implications for the risk stratification of these ED patients. That ODCAC was an independent predictor in univariate analysis but its effect was attenuated when combined with other established risk markers suggests that much of the prognostic information provided by ODCAC overlaps with that captured by clinical risk scores and cardiac biomarkers. It is clear that the high background prevalence of ODCAC in this older cohort of median age approaching 70 years is a likely explanation for the lack of discriminatory power of ODCAC in this population. This finding is not surprising, as prior studies have demonstrated similar high background rates of CAC in this age range(19). Whether a quantitative measurement of CAC burden would result in additional risk stratification beyond hsTnT and the HEART score in CP patients of this age range is a topic of further study. Of note, prior studies have further stratified ODCAC using an ordinal scoring system (mild/moderate/severe)(11). In the present study we limited ODCAC scoring to present/absent to avoid the introduction of error based on differences in imaging techniques across studies.

A notable finding of the present study is the strong NPV conferred by lack of ODCAC in our patient population that would not be considered “low risk” based on modestly elevated hsTnT levels. A prior study demonstrated a NPV of >99% for CAC =0 in ruling out obstructive CAD in CP patients of mean age 54 and with negative biomarkers (20). A meta-analysis of trials of the prognostic value of CAC=0 across 19 studies of both acute and stable CP patients with nondiagnostic biomarkers confirmed this high NPV of absence of quantitative CAC in these patients (21). The present study extends the evidence suggesting a strong NPV of negative CAC in patients with acute CP in several ways. First, prior studies used primarily the presence or absence of obstructive CAD by a CT angiography as a primary endpoint (20, 21). The present study extends the endpoint to short-term MACE, an endpoint which may not necessarily be predicted by the presence or absence of obstructive CAD. Second, while the majority of prior studies included patients with negative biomarkers (21), the present study is limited to patients with modest elevations of hsTnT, a population that is clearly at increased risk of MACE (22). Thus, it supports the concept that the NPV of lack of ODCAC exists beyond just CP patients stratified as low risk by biomarker assessment. Finally, the present study adds to the growing body of evidence supporting the retrospective use of available non-gated ODCAC for addition of CAC risk stratification (17). This latter point is worthy of further prospective studies given the attractive potential for cost and length of ED stay savings afforded by the use of “already available” patient data within crowded EDs and CP evaluation units.

Several limitations of the present study should be acknowledged. First, the retrospective design introduces potential for selection bias, both as it relates to the results and interpretation of the present study and as it relates to the application of methods of reviewing prior CT scans as part of a risk stratification strategy. This is due to the possibility that patients with available chest CT imaging may differ systematically from those without such imaging. The requirement for prior chest CT may have enriched the population for patients with comorbidities or particular healthcare utilization, potentially limiting generalizability. For example, some of the chest CTs included in this study were part of a large database of scans performed for lung cancer screening in a population of heavy prior smokers, an obvious CAD risk factor (15).

Second, qualitative coronary calcium assessment on non-gated CT scans, while practical and readily applicable, provides less detailed information than quantitative scoring. The binary classification system used in this study may not capture the full spectrum of calcium burden and its relationship to cardiovascular risk. The development of ordinal scoring or automated quantitative coronary calcium assessment methods applicable to routine chest CT could provide more detailed risk stratification information. Advances in artificial intelligence and automated image analysis may facilitate such approaches (11).

Based on our findings, several clinical recommendations can be made. First, when available, ODCAC assessment on prior chest CT imaging may provide limited supplementary risk stratification information in chest pain patients with moderately elevated troponin levels. However, based on available data this information should not replace traditional risk stratification strategies including troponin series and bioclinical risk scores. The one exception may be in patients with no ODCAC, although this finding needs to be confirmed in larger datasets and prospective trials. Clinicians should recognize that the discriminative value of coronary calcium assessment may be limited in older populations with high background rates of calcium. In such populations, calcium assessment may be most useful for ruling out significant coronary disease in the minority of patients without calcium rather than for identifying high-risk patients.

In conclusion, the present study demonstrated that qualitative coronary artery calcium assessment or ODCAC on prior chest CT independently predicted 30-day MACE in CP patients with moderately elevated hsTnT and conferred a very high NPV is this population. However, ODCAC did not provide significant incremental risk stratification value beyond hsTnT and HEART score in this study likely attributed to the high prevalence of coronary calcification in this older cohort. Future studies should evaluate the incremental benefit in younger populations with lower baseline coronary atherosclerosis burden and investigate sophisticated methods to integrate calcium assessment into ED chest pain risk algorithms.

## Data Availability

Data will be supplied upon request and approval by Geisinger Health System

## References

1. Rui P, Kang K. National Hospital Ambulatory Medical Care Survey: 2017 emergency department summary tables. National Center for Health Statistics. Available at: https://www.cdc.gov/nchs/data/nhamcs/web_tables/2017_ed_web_tables-508.pdf. Accessed January 15, 2025. DOI: 10.15620/cdc:106094

2. Collet JP, Thiele H, Barbato E, et al. 2020 ESC Guidelines for the management of acute coronary syndromes in patients presenting without persistent ST-segment elevation. Eur Heart J 2021;42(14):1289–1367. DOI: 10.1093/eurheartj/ehaa575

3. Gulati M, Levy PD, Mukherjee D, et al. 2021 AHA/ACC/ASE/CHEST/SAEM/SCCT/SCMR Guideline for the Evaluation and Diagnosis of Chest Pain. Circulation 2021;144(22):e368–e454. DOI: 10.1161/CIR.0000000000001029

4. Mueller C, Giannitsis E, Christ M, et al. Multicenter evaluation of a 0-hour/1-hour algorithm in the diagnosis of myocardial infarction with high-sensitivity cardiac troponin T. Ann Emerg Med 2016;68(1):76–87. DOI: 10.1016/j.annemergmed.2015.11.013

5. Sandoval Y, Smith SW, Sexter A, et al. Type 1 and 2 myocardial infarction and myocardial injury: clinical transition to high-sensitivity cardiac troponin I. Am J Med 2017;130(12):1431–1439. DOI: 10.1016/j.amjmed.2017.05.049

6. Six AJ, Backus BE, Kelder JC. Chest pain in the emergency room: value of the HEART score. Neth Heart J 2008;16(6):191–196. DOI: 10.1007/BF03086144

7. Backus BE, Six AJ, Kelder JC, et al. A prospective validation of the HEART score for chest pain patients at the emergency department. Int J Cardiol 2013;168(3):2153–2158. DOI: 10.1016/j.ijcard.2013.01.255

8. Vaskas A, Marshall K, Garg R, Fisher C, Bower-Stout CL, Hussain M, Matsumura ME. Effect of a heart score best practice alert on discharge decisions and outcomes of patients presenting to an emergency department with chest pain. J Emerg Med 2025;72:17–24.

9. Greenland P, Blaha MJ, Budoff MJ, Erbel R, Watson KE. Coronary calcium score and cardiovascular risk. J Am Coll Cardiol. 2018;72(4):434–447. DOI: 10.1016/j.jacc.2018.05.027

10. Arnett DK, Blumenthal RS, Albert MA, et al. 2019 AHA/ACC Primary Prevention of Cardiovascular Disease Guideline. Circulation 2019;140(11):e596–e646. DOI: 10.1161/CIR.0000000000000678

11. Foraker R, Sperling L, Bratzke L, Budoff M, et al. Opportunistic detection of coronary artery calcium on noncardiac chest computed tomography: an emerging tool for cardiovascular disease prevention: a scientific statement from the American Heart Association. Circulation 2025;152(19):e391–e401.

12. Htwe Y, Cham MD, Henschke CI, Yankelevitz DF, Shaham D. Coronary artery calcification on low-dose computed tomography: comparison of Agatston and ordinal scores. Clin Imaging 2015;39(5):799–802. DOI: 10.1016/j.clinimag.2015.04.006

13. Chiles C, Duan F, Gladish GW, et al. Association of coronary artery calcification and mortality in the national lung screening trial: a comparison of three scoring methods. Radiology 2015;276(1):82–90. DOI: 10.1148/radiol.15142062

14. Budoff MJ, Nasir K, Kinney GL, et al. Coronary artery and thoracic calcium on noncontrast thoracic CT scans: comparison of ungated and gated examinations in patients from the COPD Gene cohort. Radiology 2011;258(2):489–497. DOI: 10.1148/radiol.10100648

15. Wu MT, Yang P, Huang YL, et al. Coronary arterial calcification on low-dose ungated MDCT for lung cancer screening: concordance study with dedicated cardiac CT. Am J Roentgenol 2008;190(4):923–928. DOI: 10.2214/AJR.07.2974

16. Koh N, Nieman K. Role of cardiac imaging in acute chest pain. Br J Radiol 2023; 10.1259/bjr.20220307

17. Groen E, van Dijkman PRM, Jukema JW, Bax JJ, Hildo JL, de Graaf MA. Coronary calcifications as assessed on routine non-gated chest CT; a gatekeeper to tailor downstream additional imaging in patients with stable chest pain. IJC Heart and Vascular 2024; 52:1–7.

18. Groen RA, Jukema JW, van Dijkman PRM, Timmermans PT, Bax JJ, Lamb HJ, de Graaf MA. Evaluation of clinical applicability of coronary artery calcium assessment on non-gated chest computed tomography, compared with classic Agatston score on cardiac computed tomography. Am J Cardiol 2023;208:92–100.

19. Newman AB, Naydeck BL, Sutton-Tyrrell K, Feldman A, Edmundowicz D, Kuller LH. Coronary artery calcification in older adults to age 99: prevalence and risk factors. Circulation 2001; 104:2679–2684.

20. Grandhi GR, Mazar R, Cainzos-Achirica M, Rajan T, Latif MA, Bettencourt MS, Shaw LJ, et al. Coronary calcium to rule out obstructive coronary artery disease in patients with acute chest pain. J Am Coll Cardiol Img 2022; 15:271–280.

21. Agha AM, Pacor J, Grandhi GR, Mazar R, Khan SU, Parikh R, et al. The prognostic value of cac zero among individuals presenting with chest pain. A meta-analysis. J Am Coll Cardiol Img 2022;15:1745–1757.

22. Vigen R, Kutscher P, Fernandez F, Yu A, Bertulfo A, Hashim IA, et al. Evaluation of a novel rule-out myocardial infarction protocol incorporating high-sensitivity troponin t in a US hospital. Circulation 2018;138:2061–2063.

